# The ecology of medical care in Colombia and its relations to socioeconomic factors, 2023

**DOI:** 10.64898/2026.01.20.26344457

**Authors:** Diego A. Beltrán

**Affiliations:** Family Medicine Residency Program, Universidad El Bosque, Bogotá, Colombia

## Abstract

**Introduction:** The ecology of medical care framework describes how populations interact with different levels of health services. Despite Colombia’s formal commitment to primary health care (PHC) and near-universal insurance coverage, limited evidence exists on how health services are actually used across territories and socioeconomic contexts.

**Methods:** An ecological, cross-sectional study was conducted using national administrative data to describe health service utilization patterns in Colombia during 2023. Data were obtained from official sources, including health service provision records, health workforce registries, insurance affiliation databases, population projections, and socioeconomic indicators. Health service utilization was measured as the number of individuals per 1,000 inhabitants using different levels of care. Associations between utilization rates, socioeconomic characteristics, and physician availability were examined using negative binomial regression models with population size as an offset.

**Results:** Substantial heterogeneity in healthcare utilization was observed across municipalities and geographic regions. Most outpatient consultations were provided by non-specialist physicians, with comparatively smaller differences between primary care and specialist consultations. Emergency department visits and hospitalizations showed lower utilization rates overall, although some municipalities exhibited disproportionately high use of these services. Regions with higher poverty levels and unmet basic needs consistently showed lower utilization across most levels of care, while higher educational coverage was positively associated with specialist and inpatient care.

**Conclusions:** The ecology of medical care in Colombia reveals a health system formally oriented toward primary health care but functionally dependent on non-specialist physicians and characterized by marked territorial and socioeconomic inequities in service use. These findings suggest that universal coverage alone is insufficient to ensure equitable access and effective primary care. Understanding real-world patterns of utilization is essential to inform ongoing health system reforms aimed at strengthening PHC, improving coordination across levels of care, and addressing unmet health needs, particularly in underserved regions.

## Introduction

The concept of the *ecology of medical care* was first introduced in 1961 through the seminal article by White, Williams, and Greenberg (1) . In this classic study, the authors described patterns of health care utilization in a defined population over a specific period, illustrating how most health-related needs are addressed outside hospitals and specialized services, with primary care and informal care playing a central role. By quantifying contacts with different levels of care, the study provided a comprehensive framework to understand how health systems are actually used, rather than how they are formally designed.

Since its original publication, the ecology of medical care methodology has been widely applied in different regions and countries, including North America, Europe, and Asia. These studies have allowed health systems to better understand population-level patterns of service utilization (1–4), identify imbalances between levels of care (5–9), and explore inequities related to socioeconomic status (10–13), geographic location (2,14–16), and health system organization (4,6,7,17,18). As a result, this approach has become a valuable tool for informing health policy, planning services, and evaluating the performance of primary health care–oriented systems.

Colombia is a middle-income country located in the northwestern region of South America, characterized by marked geographic, cultural, and socioeconomic diversity. Administratively, it is divided into departments and districts, with a decentralized political structure that assigns significant responsibilities in health administration to subnational entities. These geographic and political characteristics pose important challenges for achieving equitable access to health services across urban, rural, and dispersed populations.

The country has a social security system that includes the health system as one of its core components (19). The Colombian health system is ideologically grounded in the principles of Primary Health Care (PHC) (20), offers universal population coverage, and is oriented toward guaranteeing effective access to the fundamental right to health. Over the past decades, this system has expanded insurance coverage to nearly the entire population, representing a major achievement in terms of formal access.

Despite these advances, the Colombian health system has experienced a persistent crisis related to effective access to services, quality of care, fragmentation, and financial sustainability. In response, multiple models of care have been proposed (21–23) and implemented over time, many of them explicitly oriented toward strengthening PHC as a strategy to respond to health needs across different life contexts, territories, and social conditions. However, gaps remain between the conceptual design of these models and the actual patterns of health service utilization observed in the population.

More recently, the national government has promoted a structural reform of the health system (24), seeking to transition from an insurance-based model toward a single-payer scheme. This reform aims to integrate individual and collective health actions, strengthen territorial governance, and reinforce PHC as the backbone of the system. Understanding current patterns of health service use is therefore especially relevant in this context, as it provides empirical evidence to inform ongoing policy debates and system redesign.

In this context, the present study aims to describe the use of health services in Colombia using the ecology of medical care framework and to analyze these utilization patterns in relation to multiple socioeconomic variables and indicators of health resource availability. By doing so, this study seeks to contribute to a more comprehensive understanding of how the Colombian health system is used in practice and how social and structural factors shape access to care.

## Methods

### Study design and setting

An ecological, cross-sectional study was conducted to describe health service utilization patterns in Colombia during 2023 and to examine their association with socioeconomic and health system resource factors, using the ecology of medical care framework.

### Study population and unit of analysis

The study population comprised individuals residing in Colombia who used the health system at least once in 2023. The unit of analysis was the municipality. For comparative analyses, municipalities were grouped at the departmental level or into predefined geographic regions.

### Data sources and measurement

Secondary data were obtained from official national databases: the Individual Health Service Provision Records (RIPS) analytical cubes, the Unique Registry of Human Health Talent (RETHUS), and the Single Affiliation Database (BDUA) from SISPRO; population and demographic data from the National Administrative Department of Statistics (DANE); and socioeconomic indicators from the National Planning Department (DNP). Data sources were linked using standardized municipal geographic identifiers.

### Variables

Health service utilization was measured as ratios of persons per 1000 inhabitants, using total projected municipal population as the denominator. For regression purposes, count data of health service utilization were used as a dependent variable in regression models.

Independent variables included population density, sex distribution, mean age of service users, Multidimensional Poverty Index, proportion of households with Unsatisfied Basic Needs, educational coverage (basic, secondary, and higher education), ratio of inhabitants per authorized health care institution, and health workforce availability (non-specialist physicians, primary care specialists (family medicine, internal medicine, pediatrics, gynecology/obstetrics, general surgery, psychiatry), supra-specialist physicians, and surgical supra-specialists per 10,000 inhabitants).

### Statistical analysis

Univariate analyses were conducted to describe municipality characteristics and health service utilization. Analysis of variance (ANOVA) was used to assess regional differences in independent variables and to inform selection of variables for multivariable modeling. Due to independent variable nature and non-normal distributions of dependent variables, negative binomial regression models were fitted, using the logarithmic transformation of total population as an offset variable. Analyses were performed at the municipal level.

### Software

Data extraction and organization were performed using Microsoft Excel. Jamovi and IMB SPSS v. 29 were used for univariate, ANOVA, and multivariable analyses. All *p* values lower than 0.05 were considered significant. Nested square graphics representing the ecology of medical care were produced in RStudio.

### Bias

As an ecological study based on secondary data, results are subject to ecological fallacy and potential information bias inherent to administrative databases. These limitations were considered in the interpretation of findings.

### Ethical considerations

The study used aggregated, non-identifiable secondary data. According to Colombian regulations, it constitutes research without risk and did not require ethics committee approval, in accordance with Resolution 8430 of 1993 of the Ministry of Health.

## Results

### General findings and regional distributions

Information was available for nearly all municipalities included in the analysis (Table 1). Given the skewed distribution of most continuous variables, results are presented using medians and interquartile ranges. Overall, municipalities exhibited predominantly rural characteristics, including small territorial area, low population density, moderate levels of poverty and unmet basic needs, high coverage of basic and secondary education, lower coverage of higher education, and a low density of physicians per 10,000 inhabitants.

**Table 1.**
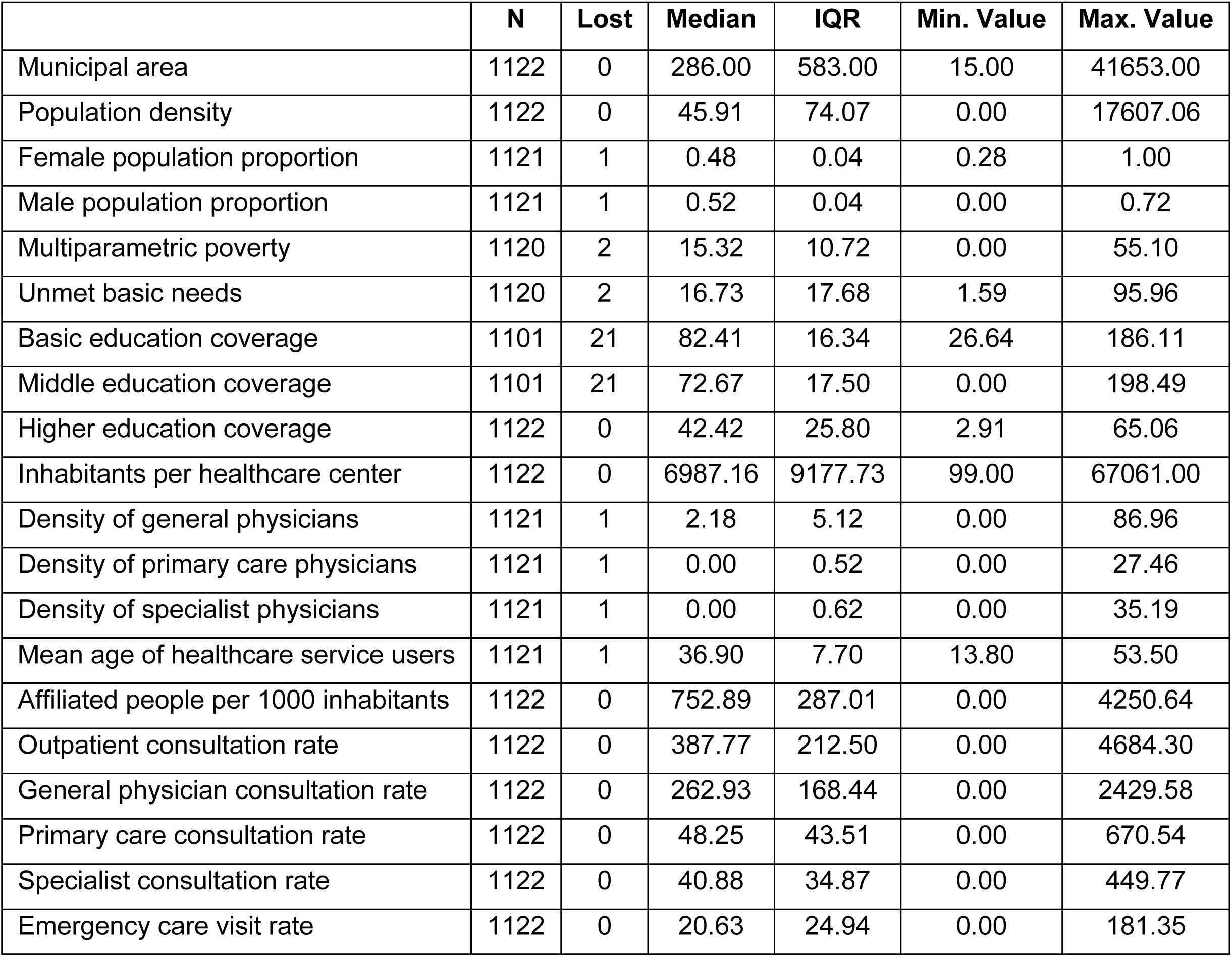
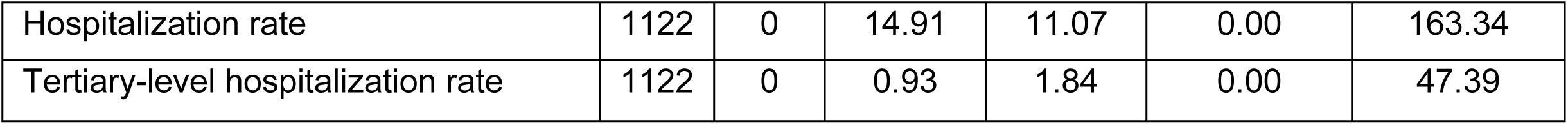
Univariate distribution of socioeconomic and healthcare usage and availability variables

As for healthcare service utilization, as shown in Fig 1, affiliation was not universal during the period of analysis. Most consultations per 1,000 inhabitants were provided by non-specialist physicians, followed by a marked reduction in care delivered by primary care physicians. However, the difference between primary care and specialist consultations was relatively small. Emergency department visits and inpatient care showed the lowest utilization rates, with limited differences between emergency visits and overall hospitalization rates, and a more pronounced gap when compared with tertiary-level hospitalizations.

**Fig 1.**
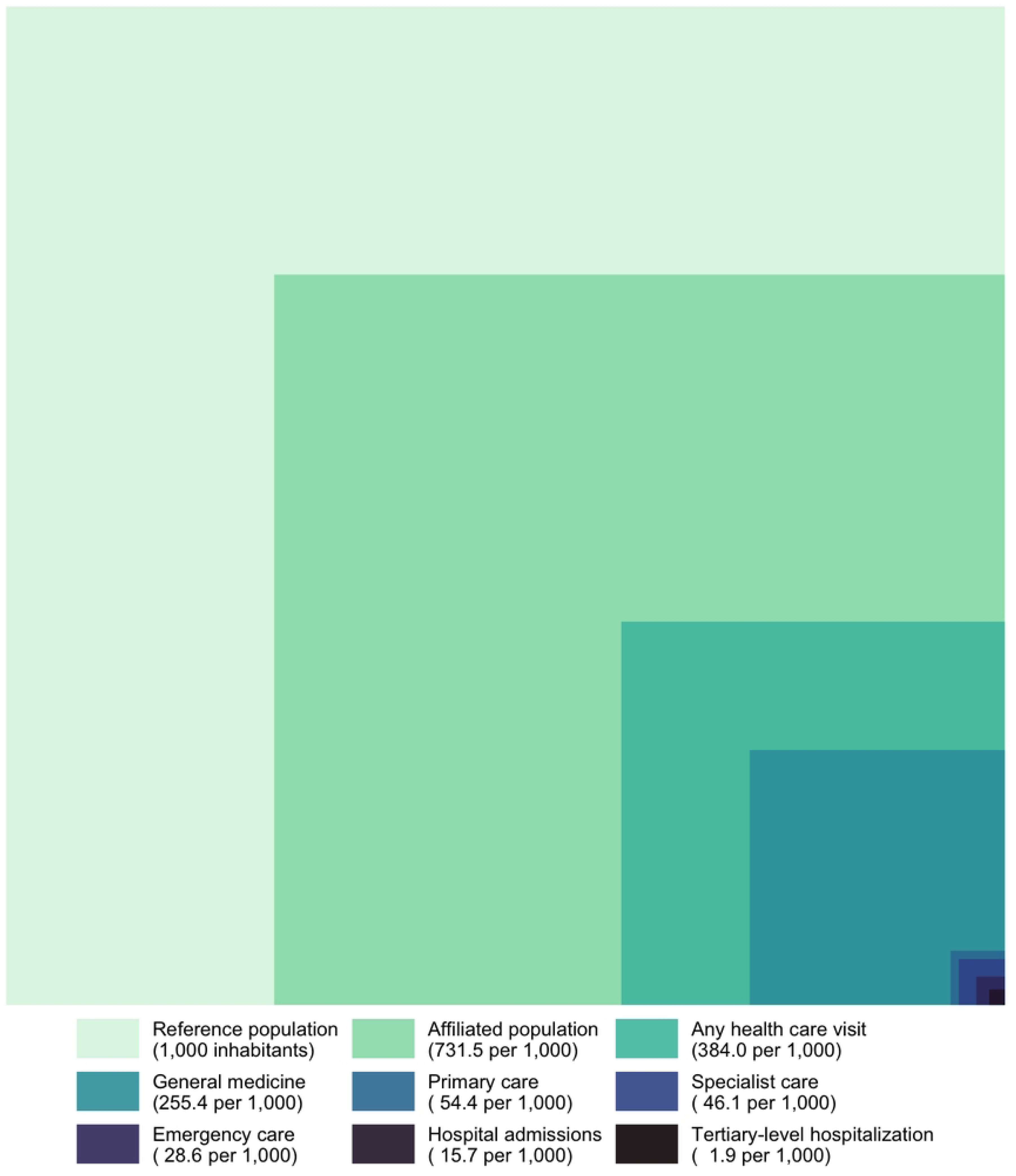
Estimated monthly number of individuals per 1,000 population who experienced a health problem and utilized medical care services across various healthcare settings

Substantial heterogeneity across municipalities was observed. Some municipalities reported no affiliated population or no consultations, whereas others exhibited affiliation or consultation rates exceeding the total resident population. Additionally, a subset of municipalities showed disproportionately high utilization of emergency and inpatient services.

All variables showed regional differences; those with greater variability are presented in Table 2. The distribution of care by regions is shown in Fig 2. The Andean region, which includes the largest number of municipalities, exhibited the highest coverage of higher education and the second lowest level of unmet basic needs, while showing intermediate levels of healthcare service utilization and availability. The Insular region, despite comprising only two municipalities, showed the highest rates of healthcare utilization and availability, together with the lowest levels of unmet basic needs and relatively low coverage of higher education. In contrast, the Amazon region exhibited the lowest levels of healthcare utilization and availability—except for emergency care visits—and the highest levels of unmet basic needs, along with the lowest coverage of higher education.

**Table 2.** Distribution of selected socioeconomic and healthcare use and availability by regions

|  | Region | N | Median | IQR | Min. Value | Max. Value |
| --- | --- | --- | --- | --- | --- | --- |
| Unmet basic needs | Amazon | 57 | 34.60 | 27.78 | 7.59 | 95.96 |
|  | Andean | 629 | 12.99 | 9.68 | 1.59 | 81.73 |
|  | Caribbean | 195 | 31.77 | 17.94 | 8.03 | 93.44 |
|  | Insular | 2 | 12.04 | 3.51 | 8.54 | 15.55 |
|  | Orinoco | 59 | 22.83 | 19.21 | 4.40 | 76.66 |
|  | Pacific | 178 | 18.55 | 21.69 | 4.11 | 83.96 |
| Higher education coverage | Amazon | 58 | 18.56 | 26.30 | 2.91 | 34.45 |
|  | Andean | 629 | 56.36 | 14.59 | 30.56 | 65.06 |
|  | Caribbean | 195 | 34.16 | 8.23 | 20.44 | 56.27 |
|  | Insular | 2 | 23.91 | 0.00 | 23.91 | 23.91 |
|  | Orinoco | 59 | 25.80 | 9.92 | 6.34 | 35.72 |
|  | Pacific | 179 | 30.83 | 5.67 | 30.06 | 44.96 |
| Density of general physicians | Amazon | 58 | 1.10 | 4.78 | 0.00 | 86.96 |
|  | Andean | 629 | 1.93 | 4.47 | 0.00 | 61.86 |
|  | Caribbean | 195 | 3.22 | 5.26 | 0.00 | 50.24 |
|  | Insular | 2 | 13.52 | 5.51 | 8.01 | 19.02 |
|  | Orinoco | 59 | 1.88 | 3.92 | 0.00 | 19.66 |
|  | Pacific | 178 | 1.86 | 5.44 | 0.00 | 56.59 |
| Outpatient consultation rate | Amazon | 58 | 224.45 | 363.66 | 0.00 | 4684.30 |
|  | Andean | 629 | 388.47 | 198.40 | 28.63 | 919.82 |
|  | Caribbean | 195 | 455.98 | 169.14 | 177.90 | 1092.95 |
|  | Insular | 2 | 373.37 | 102.17 | 271.20 | 475.53 |
|  | Orinoco | 59 | 342.39 | 305.57 | 2.29 | 951.09 |
|  | Pacific | 179 | 320.15 | 253.22 | 0.00 | 868.53 |
| General physician consultation rate | Amazon | 58 | 129.16 | 278.01 | 0.00 | 2429.58 |
|  | Andean | 629 | 273.15 | 153.30 | 14.18 | 681.91 |
|  | Caribbean | 195 | 292.43 | 143.25 | 65.50 | 762.48 |
|  | Insular | 2 | 245.05 | 70.34 | 174.71 | 315.39 |
|  | Orinoco | 59 | 218.29 | 188.15 | 1.02 | 685.85 |
|  | Pacific | 179 | 203.60 | 204.94 | 0.00 | 704.91 |
| Emergency care visit rate | Amazon | 58 | 28.26 | 42.06 | 0.00 | 147.44 |
|  | Andean | 629 | 17.18 | 19.15 | 0.00 | 143.73 |
|  | Caribbean | 195 | 46.64 | 45.93 | 5.44 | 181.35 |
|  | Insular | 2 | 32.71 | 15.72 | 16.99 | 48.43 |
|  | Orinoco | 59 | 14.81 | 16.89 | 0.00 | 97.44 |
|  | Pacific | 179 | 19.49 | 16.90 | 0.00 | 152.63 |
| Tertiary-level hospitalization rate | Amazon | 58 | 0.27 | 0.84 | 0.00 | 3.19 |
|  | Andean | 629 | 1.33 | 2.35 | 0.00 | 21.04 |
|  | Caribbean | 195 | 0.55 | 0.83 | 0.00 | 5.85 |
|  | Insular | 2 | 31.71 | 15.68 | 16.03 | 47.39 |
|  | Orinoco | 59 | 0.15 | 0.33 | 0.00 | 0.94 |
|  | Pacific | 179 | 0.71 | 1.51 | 0.00 | 14.93 |

**Fig 2.**
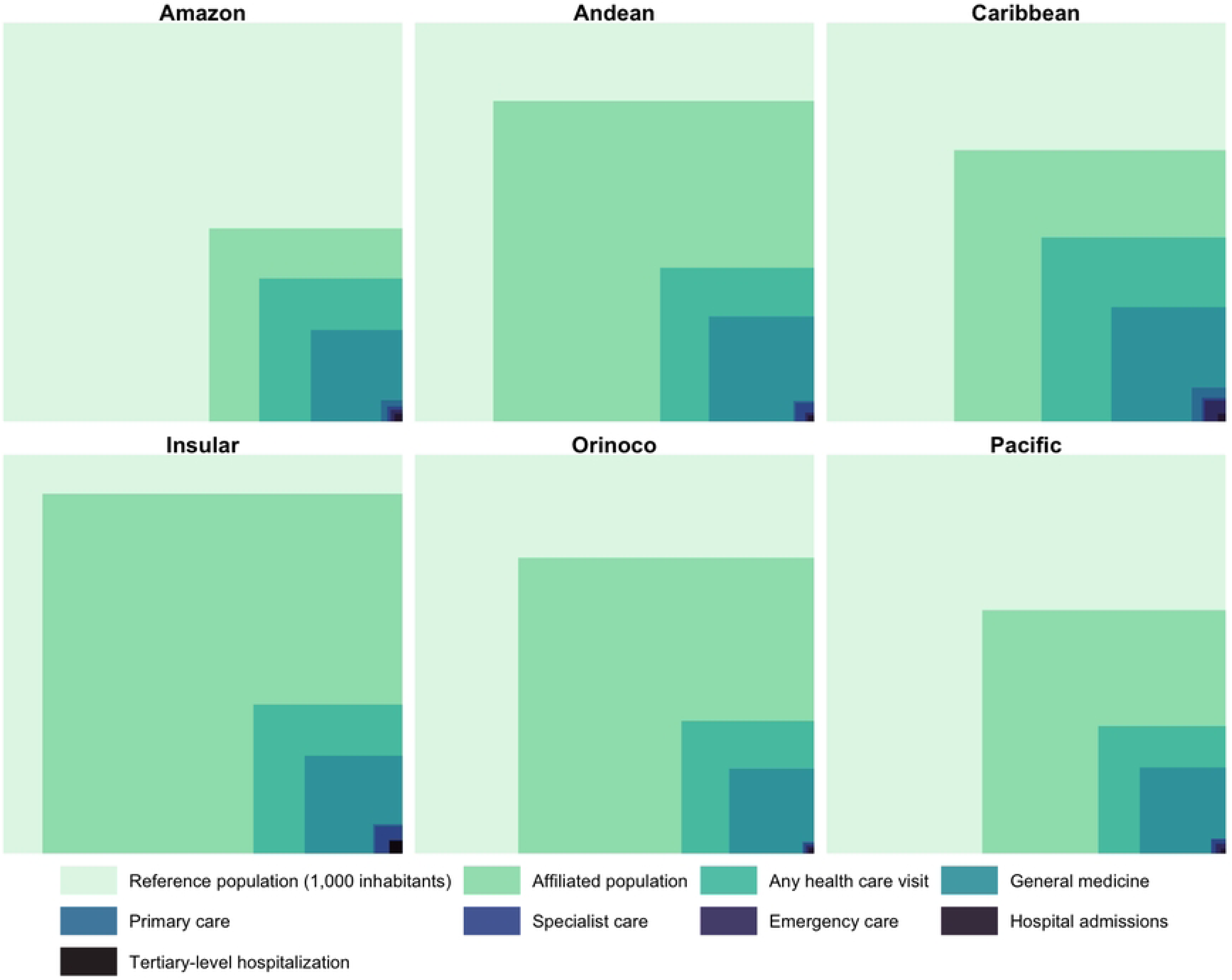
Estimated monthly number of individuals per 1,000 population who experienced a health problem and utilized medical care services across various healthcare settings by regions

To assess regional differences in key variables, Kruskal–Wallis tests were performed. Statistically significant differences across regions were observed for all variables analyzed.

### Association of socioeconomic factors and physician availability with healthcare usage rates

Marked regional differences were observed across all outcomes after adjustment for sociodemographic characteristics and healthcare availability (Table 3). Using the Andean region as the reference category, municipalities in the Insular region consistently showed the highest rates of healthcare usage, particularly for hospitalization and tertiary-level hospitalization, where incidence rate ratios were markedly higher than in all other regions. The Orinoco and Caribbean regions also showed higher rates of affiliation and outpatient consultation compared to the Andean region, although with more moderate effect sizes.

**Table 3.** Negative binomial regression model results for each level of care

|  | Affiliated people per 1000 inhabitants |  | Outpatient consultation rate |  | General physician consultation rate |  | Primary care consultation rate |  | Specialist consultation rate |  | Emergency care visit rate |  | Hospitalization rate |  | Tertiary-level hospitalization rate |  |
| --- | --- | --- | --- | --- | --- | --- | --- | --- | --- | --- | --- | --- | --- | --- | --- | --- |
|  | Exp(B) | 95% CI | Exp(B) | 95% CI | Exp(B) | 95% CI | Exp(B) | 95% CI | Exp(B) | 95% CI | Exp(B) | 95% CI | Exp(B) | 95% CI | Exp(B) | 95% CI |
| Regions |  |  |  |  |  |  |  |  |  |  |  |  |  |  |  |  |
| Andean | Reference category |  |  |  |  |  |  |  |  |  |  |  |  |  |  |  |
| Pacific | 0,95 | (0,78 - 1,15) | 0,98 | (0,81 - 1,2) | 0,97 | (0,79 - 1,19) | 0,88 | (0,72 - 1,08) | 0,96 | (0,78 - 1,17) | 1,02 | (0,83 - 1,25) | 0,86 | (0,7 - 1,05) | 1,55* | (1,24 - 1,92) |
| Orinoco | 1,18 | (0,86 - 1,62) | 1,01 | (0,74 - 1,39) | 0,94 | (0,69 - 1,29) | 0,72* | (0,52 - 0,99) | 0,75 | (0,54 - 1,04) | 0,62* | (0,45 - 0,85) | 0,84 | (0,62 - 1,15) | 0,36* | (0,25 - 0,53) |
| Insular | 1,01 | (0,14 - 7,21) | 0,93 | (0,13 - 6,65) | 0,82 | (0,11 - 5,86) | 1,31 | (0,18 - 9,41) | 1,19 | (0,16 - 8,54) | 0,57 | (0,08 - 4,11) | 1,17 | (0,16 - 8,48) | 25,02* | (3,45 - 181,41) |
| Caribbean | 1,1 | (0,9 - 1,36) | 1,32* | (1,07 - 1,63) | 1,19 | (0,95 - 1,48) | 1,96* | (1,57 - 2,44) | 1,68* | (1,35 - 2,1) | 1,73* | (1,38 - 2,16) | 1,2 | (0,96 - 1,5) | 0,78* | (0,62 - 0,98) |
| Amazon | 1,23 | (0,84 - 1,8) | 1,74* | (1,17 - 2,59) | 1,63* | (1,11 - 2,42) | 2,33* | (1,55 - 3,51) | 2,13* | (1,41 - 3,22) | 1,35 | (0,93 - 1,97) | 1,85* | (1,25 - 2,72) | 0,98 | (0,64 - 1,48) |
| Socioeconomic variables |  |  |  |  |  |  |  |  |  |  |  |  |  |  |  |  |
| Mean age of healthcare service users | 1,02 | (1 - 1,03) | 0,99 | (0,97 - 1) | 0,98 | (0,97 - 1) | 1,02* | (1 - 1,04) | 1,03* | (1,01 - 1,05) | 0,98* | (0,96 - 0,99) | 0,98* | (0,96 - 0,99) | 0,99 | (0,98 - 1,01) |
| Multiparametric poverty | 1,01 | (1 - 1,01) | 1 | (1 - 1,01) | 1 | (1 - 1,01) | 1,01* | (1 - 1,02) | 1,01* | (1 - 1,02) | 1,01 | (1 - 1,02) | 1 | (1 - 1,01) | 1,01* | (1 - 1,02) |
| Unmet basic needs | 0,99* | (0,99 - 1) | 1 | (0,99 - 1) | 1 | (0,99 - 1) | 1 | (0,99 - 1) | 0,99 | (0,99 - 1) | 1 | (0,99 - 1) | 1 | (0,99 - 1) | 0,99* | (0,99 - 1) |
| Middle education coverage | 1,01* | (1 - 1,01) | 1,01* | (1 - 1,01) | 1* | (1 - 1,01) | 1,01* | (1 - 1,01) | 1,01* | (1 - 1,01) | 1,01* | (1 - 1,01) | 1,01* | (1 - 1,01) | 1 | (1 - 1,01) |
| Higher education coverage | 1 | (1 - 1,01) | 1,01* | (1 - 1,01) | 1,01* | (1 - 1,01) | 1 | (1 - 1,01) | 1 | (1 - 1,01) | 0,99* | (0,98 - 1) | 1 | (1 - 1,01) | 1,04* | (1,04 - 1,05) |
| Density of general physicians | - | - | - | - | 1 | (0,98 - 1,03) | 1,01 | (0,99 - 1,04) | 1,02 | (0,99 - 1,04) | 1,03 | (1 - 1,05) | 1,02 | (0,99 - 1,04) | 0,97* | (0,95 - 0,99) |
| Density of primary care physicians | - | - | - | - | 1,06 | (0,94 - 1,18) | 1,07 | (0,95 - 1,2) | 1,06 | (0,94 - 1,19) | 1,05 | (0,94 - 1,18) | 1,06 | (0,94 - 1,19) | 1,09 | (0,98 - 1,22) |
| Density of specialist physicians | - | - | - | - | 0,98 | (0,93 - 1,04) | 0,97 | (0,92 - 1,03) | 0,98 | (0,93 - 1,04) | 0,94* | (0,88 - 0,99) | 0,95 | (0,9 - 1,01) | 1,01 | (0,96 - 1,07) |

\* (p < 0,05)
Notes: Dependent variable: number of affiliated inhabitants or number of inhabitants with at least one consultation during the year of interest, depending on the model. The natural logarithm of the total population was included as an offset. All negative binomial regression models showed a statistically significant improvement over the null model (likelihood ratio test, p < 0.001)

In contrast, municipalities in the Amazon region exhibited lower rates of most healthcare services, including outpatient consultation, non-specialist physicians consultation, specialist consultation, and hospitalization, with incidence rate ratios consistently below unity. An exception was emergency care utilization, which showed comparatively higher rates in this region. The Pacific region generally showed lower or similar rates of service utilization compared to the Andean region, particularly for primary care, specialist consultation, and hospitalization.

Regarding socioeconomic variables, higher levels of multidimensional poverty and unmet basic needs were associated with lower rates of healthcare usage across most models, although the magnitude of these associations was modest. In contrast, educational coverage, particularly higher education coverage, was positively associated with healthcare utilization, including specialist consultation and hospitalization rates.

In terms of healthcare supply, higher density of non-specialist physicians and primary care physicians was associated with increased rates of outpatient and non-specialist physicians consultations, while density of specialist physicians showed weaker and less consistent associations across outcomes. Emergency care and hospitalization rates appeared less sensitive to physician density, suggesting that these services may be driven more strongly by population needs and structural access factors than by local workforce availability.

## Discussion

This study applied the ecology of medical care framework to describe and analyze patterns of healthcare utilization across Colombian municipalities, incorporating socioeconomic characteristics and indicators of healthcare workforce availability. By situating these findings within a broad international body of evidence, these results contribute to understanding how a formally universal, primary health care–oriented system functions in practice within a middle-income country characterized by marked territorial, social, and geographic heterogeneity.

### Colombia within the classical and contemporary ecology of medical care

Since the seminal work by White et al. [1], the ecology of medical care has consistently demonstrated a striking stability in the proportional flow of populations through different levels of care. In the original model, approximately 750 per 1,000 adults reported symptoms in a given month, 250 consulted a physician, and only one reached a university medical center. Subsequent re-analyses in the United States have shown that, despite profound changes in medical technology, financing mechanisms, and insurance coverage, these patterns have remained remarkably stable over four decades [25,26].

These findings must be interpreted within this tradition, while acknowledging a key difference: unlike most classical ecological studies, it was not possible to quantify the base of perceived morbidity. This precludes direct estimation of the proportion of individuals with symptoms who do not seek care, a phenomenon that has been shown to vary substantially by context, culture, and access rules [8,27]. Nonetheless, the observed gradients in utilization across Colombian regions strongly suggest that unmet need and delayed care-seeking remain relevant, particularly in areas with higher poverty and structural barriers.

### Dependence on non-specialist physicians and the functional limits of primary care

A central contribution of this study is the documentation of Colombia’s heavy dependence on non-specialist physicians without formal specialty training as the main providers of outpatient care. Most consultations per 1,000 inhabitants were delivered by these physicians, far exceeding those provided by formally trained primary care or family physicians. This pattern differentiates Colombia from systems such as Israel, Canada, or Australia, where primary care is largely delivered by physicians with specific training in family medicine and supported by strong institutional gatekeeping [4,17,28].

International evidence consistently shows that the mere presence of first-contact physicians is not sufficient to ensure effective primary care. In Korea, for example, high outpatient utilization coexists with weak primary care functions and excessive reliance on tertiary hospitals for ambulatory care [5,9]. Similarly, in Austria, unregulated access and poor separation between levels of care result in high specialist use and hospitalizations, undermining system efficiency [6,7].

The Colombian pattern—where the difference between primary care and specialist consultation rates is relatively small—may reflect similar structural weaknesses. Although non-specialist physicians serve as the de facto entry point, their capacity to function as resolutive coordinators of care may be limited by training, workload, and fragmented referral pathways. This interpretation is supported by the finding that higher densities of non-specialist physicians are associated with increased outpatient utilization, while specialist density shows weaker and inconsistent associations.

### Regional heterogeneity and comparisons with rural and isolated settings

The pronounced regional heterogeneity observed in this study mirrors findings from rural and geographically isolated populations internationally. Thacker et al [2] demonstrated that the ecology of medical care is applicable to rural U.S. populations, albeit with differences in reported morbidity and utilization by race and social context. Similarly, studies in Japan’s remote islands have highlighted the critical gatekeeping role of community physicians when access to tertiary hospitals is geographically constrained [14,15].

In Colombia, the Amazon region exhibited the lowest utilization rates for most services, coupled with the highest levels of unmet basic needs and lowest educational coverage. This pattern closely resembles findings from marginalized populations in the United States, such as American Indians and Alaska Natives, where low utilization reflects structural barriers rather than lower need [16]. The relatively higher emergency care utilization in the Amazon region suggests that emergency services may function as a substitute entry point into the system when routine care is inaccessible, a pattern also observed in other underserved settings.

### Socioeconomic gradients and persistent inequities

Consistent with international evidence, socioeconomic factors in Colombia continue to shape healthcare utilization independently of formal insurance coverage. Studies from the United States, Sweden, and Belgium have shown that sociodemographic characteristics influence access and use even within publicly financed or highly accessible systems [10,12,29]. These findings align with this literature, showing that higher poverty and unmet basic needs are associated with lower utilization across most services, while higher educational coverage—particularly higher education—is associated with increased use of specialist and inpatient care.

These patterns suggest that education may act as a proxy for health literacy, navigation capacity, and empowerment in demanding services, echoing findings from Fryer et al. [11] and Hansen et al. [30], who showed that increased availability of primary care does not automatically translate into reduced hospitalizations if demand remains socially patterned.

### Absence of symptom data and implications for unmet need

A major limitation of this study is the absence of data on perceived symptoms or self-reported illness, which constitute the foundation of the ecological model.

International studies from China, Taiwan, and Hong Kong have demonstrated that the relationship between symptom perception and care-seeking is highly context-dependent, with some systems showing lower reported morbidity but high hospital dependence [3,8,31].

In Colombia, the lack of symptom data prevents direct assessment of the gap between need and utilization. This is particularly relevant given evidence from Argentina and other Latin American contexts showing that, in poorer settings, hospitalization rates may resemble those of wealthier countries while outpatient utilization remains lower [13]. These findings of lower utilization in high-poverty regions are therefore likely to underestimate true health needs.

### Strengths of a national, system-based ecological analysis

Despite these limitations, this study has important strengths. Unlike many ecological studies based on surveys or selected regions, this analysis is national in scope and relies on administrative data generated directly by the health system.

This allows for comprehensive coverage of municipalities and provides a robust picture of real-world service use, rather than self-reported behavior.

The ability to link utilization patterns with physician availability and socioeconomic indicators enhances the policy relevance of the findings, particularly in the context of Colombia’s ongoing health system reform. As demonstrated in Canada, major policy changes do not necessarily alter the underlying ecology of care if structural patterns remain unchanged [18]. This underscores the importance of aligning reforms with the actual dynamics of service use.

### Implications for primary health care reform

Taken together, these findings reinforce international evidence that strong primary health care requires more than universal coverage or high outpatient utilization.

Systems that achieve greater equity and efficiency—such as Israel or well-functioning Japanese community models—do so by integrating trained primary care physicians, effective gatekeeping, and continuity across settings [4,14,32].

In Colombia, the ecology of medical care framework highlights a critical tension: a system ideologically grounded in primary health care but operationally dependent on physicians without specialization and characterized by persistent territorial and social inequities. By revealing how services are actually used across regions and populations, this study provides empirical evidence to inform reforms aimed at strengthening primary care capacity, improving coordination, and addressing unmet need—particularly in underserved territories.

## Data Availability

All relevant data are within the manuscript and its Supporting Information files.

## Acknowledgements

The author would like to acknowledge the Colombian Ministry of Health and Social Protection and the institutions responsible for the generation and maintenance of the administrative health databases used in this study, including SISPRO, DANE, and the National Planning Department. The author also thanks all health care workers and administrative personnel whose routine work in data collection and reporting made this analysis possible. The views expressed in this article are those of the author and do not necessarily reflect the official positions of the institutions involved.

